# Navigating HIV Self-Testing: concerns among adolescents and young people aged 15-24 years in Uganda

**DOI:** 10.1101/2025.08.11.25332607

**Authors:** Richard Muhumuza, Denis Ndekezi, Rehema Nagawa, Esther Awino, Chiti Bwalya, Madalitso Mbewe, Joanita Nassali, Jackiline Namara, Felix Rutaro, Musonda Simwinga, Virginia Bond, Janet Seeley, Andrew Sentoogo Ssemata

## Abstract

**Introduction:** HIV self-testing (HIVST) has the potential to overcome barriers to conventional clinic-based HIV testing services by offering a convenient, private, and confidential way to test. This study aimed to explore the concerns about HIV self-testing among adolescents and young people (AYP) aged 15-24 years in Uganda, where HIV self-testing is still not widely available.

**Methods:** We conducted 14 audio-recorded in-depth interviews and six focus group discussions with adolescents and young people in Wakiso district, Uganda, between March 2021 to February 2022. These interviews were transcribed verbatim and analysed thematically using the socio- ecological model. All participants provided written informed consent and assent before participating in the study.

**Results:** AYP viewed HIVST as a potentially helpful and acceptable testing method. However, several concerns emerged. At the individual level, participants expressed fear of suicidal thoughts if one tested HIV positive, lack of adequate information, increased risky sexual behaviours, neglect of other HIV preventive measures, and misinterpretation of test kit results. Interpersonal concerns were centred on partner violence, parental coercion, and social rejection. At the community level, participants noted the potential for stigma, unintended pregnancies, and discrimination. Institutional concerns focused on the lack of referral services and inadequate counselling following HIV self-testing. At the structural level, limited accessibility for persons with disabilities was a key concern.

**Conclusion:** While AYP have established that HIVST is essential, various concerns need to be addressed to improve its acceptability, take-up and utilisation. Providing more explicit information about the testing procedure, clarifying any misinterpretation of results, and providing easy access to counselling services, especially at distribution points, is crucial towards guaranteeing the effective roll-out of HIVST among young people. Special attention must also be given to marginalised populations, including people living with disabilities, to ensure fairness in their access and utilisation of services. Implementation of all this will be crucial towards maximising the potential benefits of HIVST in Uganda’s HIV prevention efforts.

## Background

The HIV epidemic remains a public health concern worldwide, with 37.0 million people living with HIV, 1.3 million new HIV infections, and 630,000 AIDS-related deaths in 2024 (UNAIDS, 2025). In 2021, adolescent girls and young women (AGYW) represented 63% of all new HIV infections among young people in the region, despite being only about 10% of the population. Among adolescents, 80% of new infections are in girls aged 15–19 years, and AGYW are twice as likely to be living with HIV as young men of the same age group (Murewanhema, Musuka, Moyo, Moyo, & Dzinamarira, 2022; UNAIDS, 2021).

Many HIV preventive methods have been advanced that include the Abstinence, Being faithful and Condom use (ABC) strategy, Pre-Exposure Prophylaxis (PrEP), Post-Exposure Prophylaxis (PEP) and HIVST, but despite these, HIV infections continue to increase (van Sighem & van der Valk, 2022). HIV self-testing is a process whereby a person who wants to know his or her HIV status collects a specimen, performs a test and interprets the test result in private (Hector et al., 2018; Matovu et al., 2020; A. UNAIDS, 2014; Wilson et al., 2022).

Studies have explored HIV self-testing (HIVST) among the general and key populations across the globe, including sex workers, fisherfolk, and men who have sex with men (MSM) (Burke et al., 2017; Bwalya et al., 2020; Mulubwa et al., 2019; K. Ortblad et al., 2019; Tucker, Wei, Pendse, & Lo, 2015; Zeleke, Stephens, Gesesew, Gello, & Ziersch, 2024). These studies found that HIVST was highly acceptable, although with low levels of knowledge about the testing process.

Although HIVST was found to be acceptable, several concerns have been reported, including increased stigma, particularly among adolescents, perceptions by communities that self-testing may lead to unintended consequences, such as social isolation, anxiety, and depression (Hlongwa, Mashamba-Thompson, Makhunga, Muraraneza, & Hlongwana, 2020; Indravudh et al., 2017; Jamil et al., 2021; Napierala Mavedzenge, Baggaley, & Corbett, 2013).

Additionally, HIVST has been associated with violence, particularly among vulnerable populations such as women, sex workers, and MSM in settings where gender-based power imbalances are prevalent (Kumwenda et al., 2019; Schaffer, Agot, & Thirumurthy, 2017). In some contexts, partners may react violently if an unexpected result is disclosed, or HIVST may be coercively introduced in relationships (Bulterys et al., 2023; Naughton et al., 2023).

While there has been a high uptake and coverage of HIV self-testing (HIVST), particularly among young adults, highlighting its effectiveness in reaching underserved groups such as men, young people, and first-time testers Despite promising evidence showing high uptake and coverage of HIVST among young adults especially in reaching underserved populations such as men, first-time testers, and young people (Hatzold et al., 2019; Sharma, Ying, Tarr, & Barnabas, 2015; Tonen-Wolyec et al., 2019) access to HIVST among AYP remains uneven and suboptimal.

Additionally, there is a limited understanding of how AYP aged 15–24 years navigate the health system to access and use HIVST effectively. Existing studies have highlighted key challenges in this age group, including failure to interpret test results and the absence of counselling following the test among adolescents (Tonen-Wolyec et al., 2019; Zeleke et al., 2024), and linkage to care (Phongphiew et al., 2021). In the Ugandan context, additional barriers such as health facilities being perceived as confusing, physically distant, unaffordable, and staffed by older, judgmental healthcare providers further deter AYP from seeking HIV testing services (Kalibbala et al., 2022).

In Uganda, HIVST was initially adopted in 2018 as a testing approach for Key Populations (K. Ortblad et al., 2017; World Health Organisation, 2019). HIV self-testing kits were distributed through pilot projects. The Ugandan government, with support from implementing partners, have made significant investments in HIVST services (Nsereko et al., 2024). HIVST kits have been made available for sale in pharmacies, but not in public facilities (Nasuuna et al., 2022). However, uptake and utility by adolescents and young people in Uganda remain suboptimal, and this is likely to affect the attainment of the 95-95-95 UNAIDS goals to end HIV by 2030.

Given that HIV testing remains a critical entry point for both prevention and treatment services, understanding the concerns AYP face around HIVST is essential. This study aimed to explore HIVST concerns among adolescents and young people aged 15-24 years in Uganda before the HIVST rollout. The insights generated will inform more youth-responsive HIVST strategies and contribute to improving testing uptake and outcomes among this priority population.

### Theoretical orientation

We adopt the social-ecological model - SEM (Bronfenbrenner, 1994; Kaufman, Cornish, Zimmerman, & Johnson, 2014) to explore concerns of AYP about HIV Self-testing. The SEM model proposes that an individual’s health behaviour is influenced by multi-level, interdependent factors, including individual, interpersonal, community, and broad society or system levels (Kilanowski, 2017).

## Methods

### Description of the study setting and Study design

Using a case study design of three communities, this study was nested within another large exploratory mixed methods study that aimed to investigate the knowledge, acceptability, and social implications of a peer-to-peer HIV self-testing (HIVST) distribution model among adolescents and young people aged 15–24 years in Uganda. The study was implemented in three fishing landing sites of Kigungu, Gerenge and Nakiwogo in Wakiso district between March 2021 and February 2022. These fishing communities have a vibrant young population engaged in fishing activities and the associated hospitality industry (bars, restaurants and lodges) (Sileo, Kintu, Chanes-Mora, & Kiene, 2016).

### Study population

The primary population for the HISTAZU study was male and female AYP, aged between 15 and 24 years, residing in areas selected from the 3 study sites in Wakiso district.

### Selection and recruitment

Participants were purposively selected through the village information meetings and peer mobilisation activities with the support of Community Health Extension Workers (CHEWs), formerly Village Health Teams (VHTs). The CHEWs mobilised young people to attend community meetings where the study team provided information about the study. Those interested in participating in the study were then invited to participate and then later registered and later enrolled on the study.

### Data collection

Data for this study was collected through Focus Group Discussions (FGDs, n= 6) where 60 AYP participated. Each FGD was conducted by two researchers: one moderating the discussion with the other observing and taking notes of the proceedings. The FGDs were a useful method for collecting general community perspectives and shared experiences related to HIVST.

To yield additional detailed insights, key themes explored in the FGDs were further explored through the in-depth interviews IDIs (n=14) with the gaol of obtaining detailed narratives of AYP’s views on HIVST as some AYP perceived the topic of HIV and testing as sensitive.

These FGDs and IDIs explored AYPs perceptions of as well as preference of HIVST distribution models, points of access, and type of test kit (blood-based or oral). They further explored perception of and experience with barriers and facilitators of access to and use of HIVST among AYP in Uganda.

Utilising both FGDs and IDIs enabled the researchers to navigate a wide variety of different views about a particular issue and provided the opportunity for the researcher to observe how individuals collectively made sense of HIV self-testing and the meanings they attached (Bryman, 2016).

### Data management and analysis

All the IDIs and FGDs were audio-recorded, transcribed verbatim and translated into English (for those in Luganda, a local dialect). The transcripts, alongside the audio recordings and notes taken during the data collection, were reviewed to ensure consistency and that meaning was not lost during the transcription and translation. The data was uploaded onto the server for secure storage.

Using a thematic data analysis approach and guided by the SEM, all parts of the data transcripts and notes from observations were manually open coded to identify possible themes based on the study objectives and on new themes which came from the data collected. Transcripts were read several times to ensure the context of the data was fully understood. Themes emerging from the data were then written up to identity individual, household or interpersonal, community and broad society or system levels factors that influenced experiences of AYPs with HIVST in the study population

### Positionality statement

This work was carried out by a team of experienced Zambian, British and Ugandan researchers. All members of the research team have experience working with HIV programs in communities. Zambian researchers are social scientists with prior qualitative research experience in HIV prevention and treatment and are fluent in English. British and Ugandan researchers who have years of experience in collecting qualitative data in Ugandan community settings.

During data collection and analysis, reflexive practices and member checking strategies were used to improve the rigour of the work. Researcher qualifications, social positions, and work experience were considered together with researchers’ respective backgrounds, beliefs, and assumptions and how these may have influenced the analysis and interpretation of study results. Weekly meetings were used for peer debriefing and comparison of interpretation results among the Uganda team. These results were then shared with the Zambian team for review. These steps increased the trustworthiness and credibility of this research, ultimately contributing to a more comprehensive understanding of how AYPs in Uganda experienced HIVST.

### Ethical considerations

This study received approval from the Uganda Virus Research Institute Research and Ethics Committee (GC/127/20/05/767), the Uganda National Council for Science and Technology (SS446ES), and the London School of Hygiene and Tropical Medicine (Ethics ref 22588). Participants above 18 years provided written informed consent, while written parental consent and assent were obtained from those below 18 years before participating in any study procedures.

## Findings

### Socio-demographic characteristics

A total of 82 participants took part in both the in-depth interviews and group discussions, primarily adolescents and young adults aged 15–24, with the largest proportion falling in the 19–22 age group. The demographics are presented in Table 1.

**Table 1:**
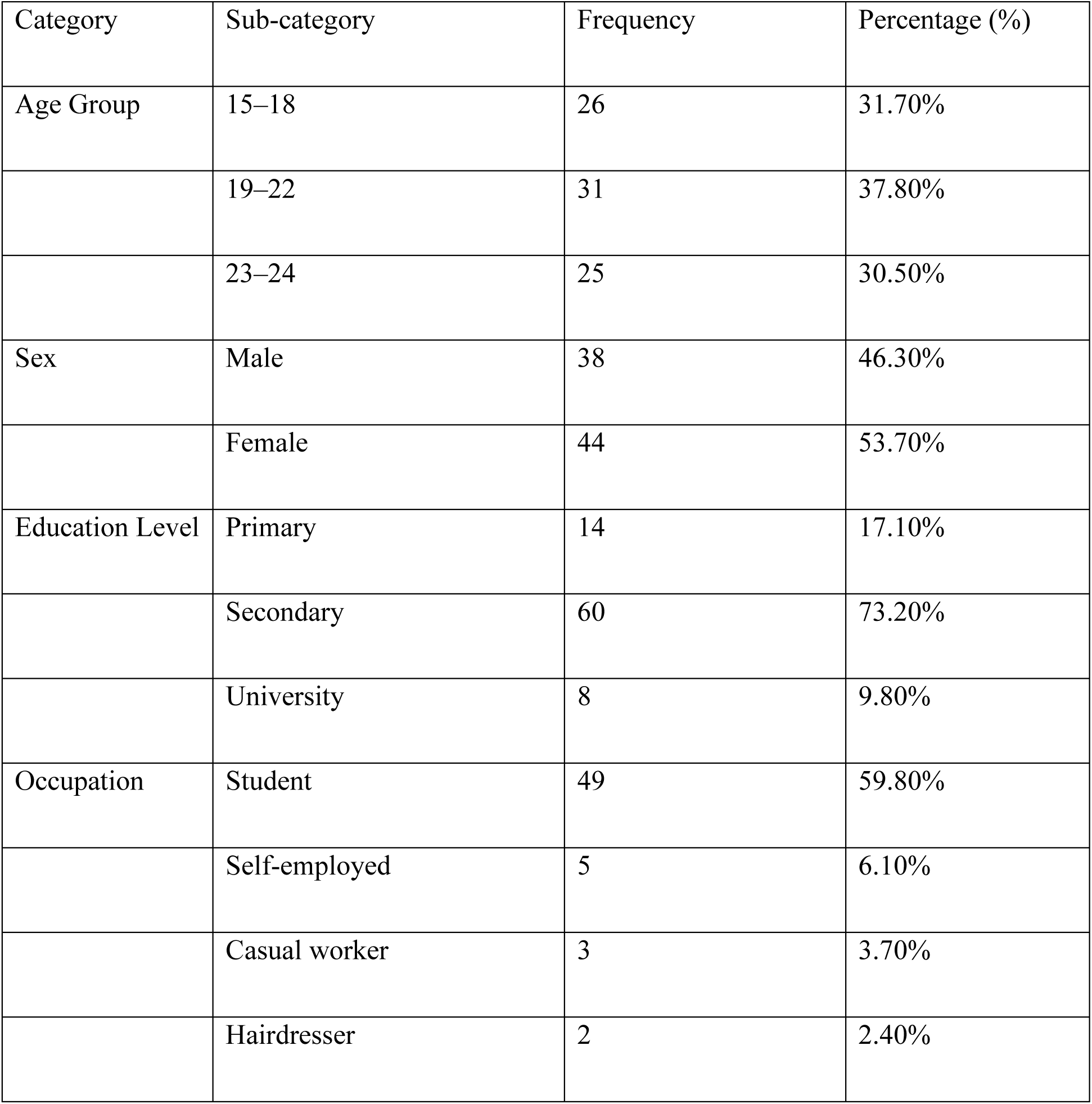
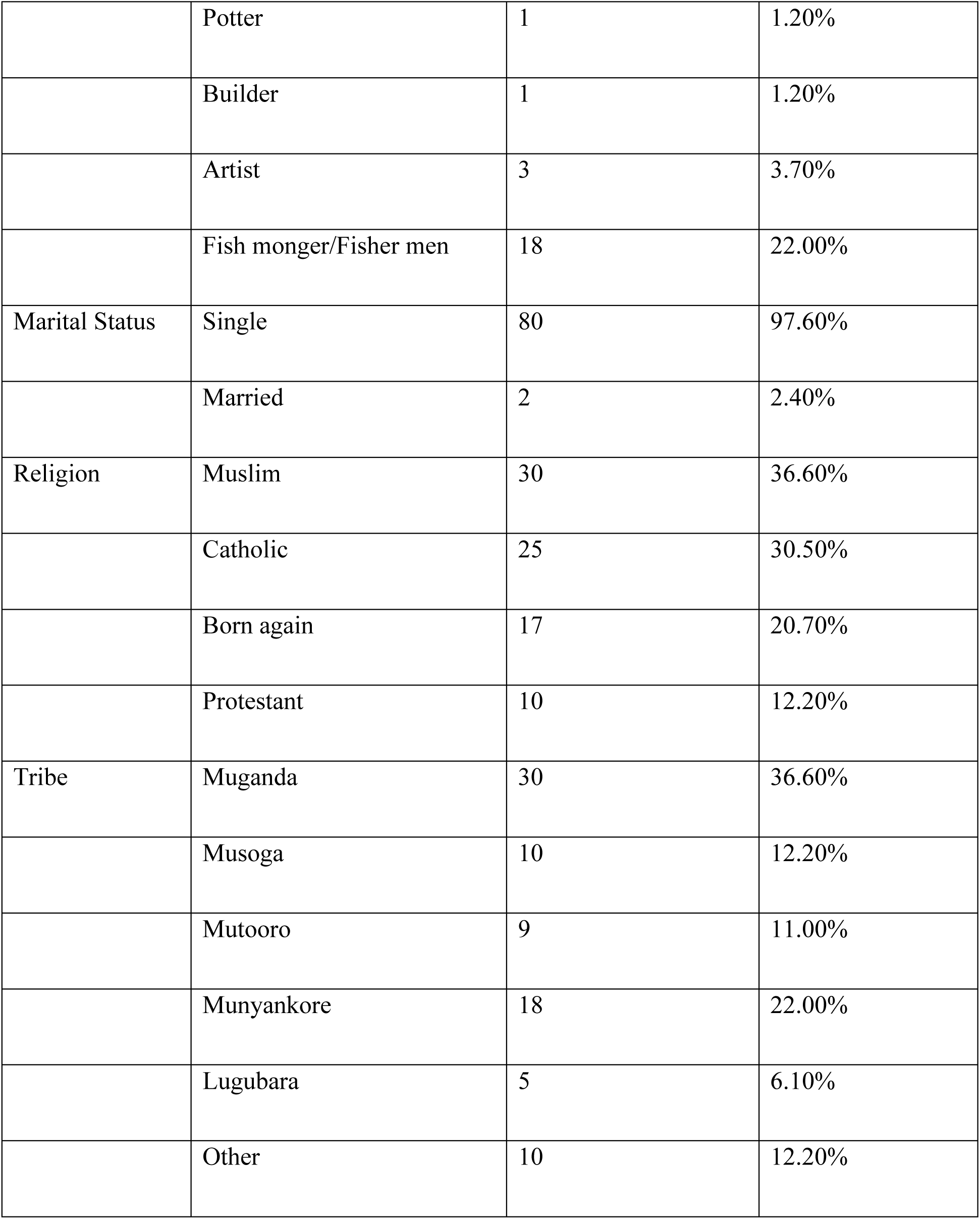
Participant demographic characteristics.

During the analysis, we identified key themes describing concerns among adolescents and young people. These were categorised into five levels (individual, interpersonal, community, institutional and structural) based on the socio ecological model constructs.

### Individual-level concerns with HIV Self-Testing

The AYP concerns about self-testing at the individual level included anticipated social harm as a result of self-testing, failure to link with care and treatment, increased sexual behaviour among AYP, and misleading results due to failure to interpret and not being inclusive of other special needs people.

Participants narrated that there were social harms perceived to follow HIV self-testing, especially if the results were positive. Without adequate emotional support, positive results may trigger intense psychological distress, including suicidal thoughts, particularly among vulnerable youth as one noted below.

> *“In case results are out and one has not received enough counselling, some people develop fear and lose hope, and in the end, they end up thinking of committing suicide because they see no meaning in life. They must receive enough counselling to accept the situation.” IDI Male 20-25 years*

Participants who would obtain a positive HIV diagnosis through self-testing thought that they would experience emotional distress and suicidal tendencies in the immediate aftermath of testing. This was attributed to the absence of pre- and post-testing counselling. This exacerbates sentiments of despair and psychological disorientation.

> *“That is also not good. It has its disadvantages. You can test yourself and turn positive and, in the process, commit suicide. So, you need to carry out the test with someone who is an expert and can counsel you in case of anything.” IDI Male 15-20 years*

### Changes in sexual lifestyle

The majority of participants anticipated that HIV self-testing would increase risky sexual behaviour among their peers, particularly unprotected intercourse. This worry stemmed from the assumption that AYPs who test HIV-negative would engage in more casual and unprotected sexual encounters, potentially leading to increased risky sexual behaviour.

> *“Sexual activity and risk-taking among adolescents will increase. As you said, once adolescents know their sexual partner’s HIV status is negative, they will engage in unprotected sexual intercourse.” IDI Male 15-20 years*

Additionally, some participants emphasised the importance of integrating risk reduction messaging into HIV self-testing programs. Similarly, providing accessible sexual health services and monitoring the impact of self-testing on adolescent sexual behaviour would be critical in preventing such unintended consequences, as one participant narrated.

> *“I think they should teach us more about HIV self-testing and prevention when they give us these self-testing kits. If I test negative, I don’t want to get complacent and forget to use condoms. We need to know how to protect ourselves and our partners.” GD Female 20-25 years.*

Other AYP considered HIVST as a source of empowerment, creating a sense of positive control over one’s sexual lifestyle and health, especially in times when one feels at heightened risk of HIV.

> *“Most cases when adolescents know they are HIV negative, sexual behaviour also reduces because they want to remain HIV negative. They do not over engage in sexual intercourse because they know that when they do engage in sexual intercourse they are at risk of contracting HIV. In most case when adolescent sexual partners know each other’s status, it increases faithfulness and trust in that couple.” IDI Male 20-25 years*

Some of the participants further expressed that HIVST will provide an opportunity for young people to adopt more responsible sexual behaviour responses and preventative lifestyles, viewing the HIVST model as an additional protector.

### Inability to interpret test results

Participants expressed concern about HIVST regarding its reliability and usability. For example, some of the participants noted that incorrect use or interpretation of HIVST could yield false negative results, leading individuals to forego linkage to care and antiretroviral therapy.

> *“HIVST can mislead some people, especially when used incorrectly and produce wrong results that they are HIV negative, yet they are HIV positive, thus end up not linking to care to get medication.” GD Males 15-20 years*

> *“For someone who doesn’t understand how to use the kit, when it reaches T and gets stuck, he or she might not understand whether it is positive or negative.” GD Females 15-20 years*

Furthermore, some participants, particularly those with limited literacy, expressed scepticism about HIVST due to difficulties in understanding test results, opting instead for standard clinic based rapid testing conducted by healthcare professionals.

> *“Some adolescents will prefer to use the standard test because they do not know how to read or understand the results after testing and thus do not trust the self-testing kit.” IDI Male 20-25 years*

Beyond misinterpretation of results, participants raised concerns about the denial of the positive test. This was linked to the premise that some AYP would not be able to believe in the test results from the self-testing kits. For many AYP, blood-based HIV testing is what was trusted to provide accurate test results, as HIV was commonly assessed through blood and not saliva. additionally, they found it unusual that an oral saliva-based swab test could be used to test for a virus found in the blood.

> *“Some of us find it a challenge to accept the truth. You might turn positive and think that the kit was faulty. Other young people will ask you if you have ever seen HIV/AIDS tested through saliva or if HIV/AIDS is transmitted through saliva because the kit does not use blood during testing.” IDI Male 15-20 Years*

### Neglect of other HIV preventive methods

Participants mentioned HIVST would offer relief from the discomfort of using condoms to protect themselves against HIV after testing negative with the kit. This meant that they were more likely to abandon other protective measures. AYP also noted that when one tests with his partner using the kits and both get a negative result, they find no reason to engage in sexual intercourse with a condom.

> *“The kits may decrease use of condoms because people who test negative will have unprotected sex thinking that they are HIV negative and end up acquiring other infections. GD Males” 20-25 Years*

Some participants recognised that HIVST Kits, unlike condoms, would not prevent Sexually Transmitted Infections (STIs). This pointed to the fact that condoms were used to prevent other related risks such as STIs. Those who tested negative would be influenced to engage in sex without any other protection, thereby increasing their risk of contracting STIs.

> *“What I can tell you is that there is going to be an increase in infection rates related to STDs. Apart from HIV, these other STIs are going to increase because most adolescents’ concerns are about HIV it is not all about HIV. Once they test Negative, they are going to have unprotected sex, and this will put them at high risk of contracting these STDs.” GD Females 20-25 Years*

Similarly, unprotected sex would lead to increased cases of unintended pregnancies among adolescents and young people. Some participants argued that after testing negative, they would find no need to use condoms, leading to unintended pregnancies, as the following excerpts reveal.

> *“After testing with your partner and finding out that you are HIV negative, there is no need to wear a condom. But the problem is that it is going to cause Pregnancies. GD Males 20-25 years*

> *There will be an Increase in unwanted teenage pregnancy. After self-testing and knowing that, they are both HIV negative. They will not use condoms; what will happen next will be an unplanned pregnancy.” IDI Male 15-20 Years*

The unplanned pregnancies resulting from unprotected sex would lead to increased abortion rates, as many young girls are unprepared for pregnancy and family responsibilities.

> *“As a result of unplanned or unintended pregnancies, AYP stressed that this would increase the rate of abortions among many young girls who are not ready to carry their babies. “Since they are going to have unprotected sex the rate of abortion is going to increase because most of these girls don’t want pregnancy and are not ready for family issues” GD Females 20-25 Years.*

### Interpersonal level concerns

At the interpersonal level, the AYP narrated that those individuals who had not disclosed their status were concerned about how their partner would react if they were to learn of their HIV status, fearing rejection and violence against them. HIVST was therefore viewed as a bad idea for couples to test themselves without any supervision and counselling.

> *“Self-testing will cause domestic violence in homes. For example, among married couples, self-test for HIV/AIDS with your partner, and you are the only two in the house. In addition, the result comes out, one is positive, and another is negative. At that moment, a man can physically abuse the woman and even separate without getting proper counselling.” IDI Female 15-20 years*

> *“Self-testing is going to separate couples. From what I see is your partner will bring the test kit and want you to test together. Yet you are HIV positive, but you lied you are negative. Then you will regret why they invented self-testing kits. Because of this, relationships are going to break.” IDI Female 15-20 years*

The sudden revelation of your HIV status may frequently result in relationship breakdowns underscoring the need for comprehensive counselling and support services while using HIVST to mitigate conflict risks.

Additionally, some of the participants expressed concerns that HIVST kits would facilitate coercive testing by parents, compromising their autonomy and privacy, and undermining their ability to make independent decisions about their health.

> *“Parents will start bringing them home and force us to test for HIV” GD Female 15- 20.*

While some worried about parents forcibly administering home tests, others anxiously contemplated explaining positive results, anticipating parental judgement and overreactions.

> *“In case I turn positive, what will I explain to my parents? They might think about many things,” IDI female 15-20 years.*

This dual concern highlights the delicate dynamics of family relationships, HIV stigma, and AYPs’ vulnerability, underscoring the need for targeted support and guidance on disclosure, confidentiality for the parents and young people.

## Institutional-level concerns

### Absence of counselling and linkage to care

The participants in our study revealed sentiments about the absence of counselling and linkage to care challenges among adolescents who would discover their status through the HIV self-testing option. This was discussed in three aspects: absence of counselling, absence of referral services for confirmatory testing and failure to link to care and further treatment, as well as fear of picking medication from the service providers.

The majority of the participants highlighted HIVST as an important HIV testing strategy for AYP; however, the lack of counselling services if an individual tested positive creates fears and concerns in the uptake of the HIVST model.

> *“HIVST would be good only that it lacks counselling which is very important because once you discover that you are positive, you can end up in deep thoughts [anxious] unlike when you access counselling where you can be helped and guided on how to start medication so it will be hard to use the HIVST method” FGD Male 20-25 years.*

Participants believed that a person must be prepared to receive the outcome from the results of the HIVST kit. The participants felt that they would have nobody to reach out to, which was seen as dangerous, as somebody would collapse, engage in risky sexual behaviours and get depressed because of a positive HIV self-test.

> *“Some people may struggle to cope after self-testing and may turn to alcohol. For such individuals, self-testing may not be appropriate. At the hospital, when someone is unwell, they receive counselling and emotional support to ensure they begin ART. But with self-testing, there is no one to offer comfort or guidance you are left to cry alone until it becomes too much…” IDI Male 20-25 years.*

> *“Yes, with self-testing, there is no counselling… adolescents do not know many things. We still need counselling to be informed of the current information concerning HIV/AIDS so that we don’t live the same lifestyles. In this current era, we need to be ready for everything.” IDI male 15-20 years*

Referral and linkage for confirmatory testing was viewed as another concern. Some of the participants revealed a fear of going to the facility for a confirmatory test. It would therefore take a long process of convincing somebody to go for a confirmatory test.

> *“Some partners fear going to the hospital and because of that, I can try my level best to convince her to test. I can also tell her that in case she didn’t understand, she can go to the health worker for more clarity, and we can try it out. So, in the process of testing, I also tell her that there is something I didn’t understand and convince her that we go for a confirmatory test at the health facility.” IDI Male 20-25 years*

> *“That is what I was telling you if someone tests HIV positive, he or she will fear to come to the health facility to confirm her or his results. “Why would I go to the hospital, yet I already know my results”. Should I go to the hospital, and they give me all that medication which is supposed to be taken every day!!” IDI Male 20-25 Years*

Integrating self-testing with comprehensive support, training healthcare providers in adolescent-friendly care, and developing targeted interventions can help ensure timely linkage to antiretroviral therapy and improve health outcomes among adolescents living with HIV.

### Structural level concerns Complexities of the use of HIVST

The HIV self-testing kits were viewed to pose significant accessibility challenges for visually impaired individuals, particularly the blind, who face difficulties in reading and interpreting results and lack autonomy in testing. AYP noted that these current kits require visual interpretation, excluding those who cannot see, and reliance on others for result interpretation compromises confidentiality and increases stigma.

> *“This model is going to be hard for the blind because she can collect the samples very well but cannot interpret the results.” GD Females 15-20 Years*

> *“A blind person can’t read the result because even if he tests himself, how is he going to interpret the results? Yes, it is not good for blind people. IDI Female 20-25 years*

> *It is true. The self-test kit is not user-friendly for the blind because there is no way a blind will know whether he is positive or negative.” IDI Female 15-20 years*

These findings not only highlight the psychological risks associated with receiving a positive result through HIV self-testing but also reveal critical gaps in the accessibility of self-testing kits for certain populations.

## Discussion

HIVST is a new approach to HIV testing and was rolled out in Uganda in 2018 among adults (Bbuye et al., 2022) but at the time of our study, HIVST had not been rolled out to the general population. We, therefore, set out to understand adolescents and young people’s concerns about HIVST once rolled to the entire population. Despite UNAIDS recommendation for self-testing across all age groups, over 70% of adolescents and young people were unaware of HIV self-testing (UNAIDS, 2021). Both older and younger adolescents had similar concerns and what came out most was the feeling of self-harm if one tested positive. This means that the participants were more likely to be depressed and consequently commit suicide or even harm the person they anticipate having infected them with HIV. This was also linked to non-disclosure and consequently failure to link to care which contravenes the 95-95-95 as stipulated by UNAIDS and World Health Organisation.

The anticipation of social harm among adolescents and young people in our study was consistent with results from other populations (Burke et al., 2017; Obiezu-Umeh et al., 2021) who found that people feared that there was a likelihood of a person committing suicide upon learning of positive results and lack of counselling.

Our findings support a review by Brown, Djimeu, and Cameron (2014) that highlighted concerns about potential unintended harm, including psychological harm when testing and counselling are decoupled, social harm from the potential unethical use of HIV self-test kits or a nonreactive (negative) HIV self-test resulting in justification for unprotected sex, and medical harm from greater potential for inaccurate results. It is imperative to note however that from the review articles, the potential concern of suicide with HIV self-testing was mentioned, but no evidence of suicide after an HIV self-test was reported (Brown et al., 2014). On the other hand, a systematic review by (Johnson et al., 2017) revealed that large-scale trials and implementation of HIVST have yet to identify any reports of suicide or self-harm following HIVST. Other issues reported by studies conducted in Kenya, Zambia and Uganda reflected that participants there were concerned with the fact that HIV self-testing would breed gender-based violence as well as mistrust and suspicions among couples (Chanda et al., 2017; Kumwenda et al., 2019; Masters et al., 2016).

The absence of counselling, referral and linkage to care reported in our study where HIVST has not been rolled out to the general population continues to highlight the need to examine and evaluate the place of HIV pre-test and post-test counselling. From the findings, the concerns about anticipated social harms and an increase in risky sexual behaviour are more definite when testing occurs in the absence of counselling (Brown et al., 2014).

Our findings affirm that additional research is needed to develop innovative and effective mechanisms to inform and support those using HIV ST to seek follow-up confirmatory testing services, promote linkage to counselling and HIV reporting, care and partner notification (Johnson et al., 2017; Spielberg, Levine, & Weaver, 2004).

With the great potential to reach individuals who have never tested and otherwise may not report to a health facility for testing, HIVST is key to many prevention interventions including behaviour change communications to reduce risky behaviour (Brown et al., 2014). However, linkage to care was a contested issue where the test result came out positive. Like what other scholars have found there was low linkage to care between men who have sex with men and transgender women (Bustamante et al., 2017). Some of the reasons for low linkage to care were attributed to the failure to evaluate the number of people who self-test for HIV as well as the lack of counselling (Njau et al., 2020; Tucker et al., 2015; Zanolini et al., 2018). Another study also revealed that HIV self-testing removes the opportunity for clinical testing to evaluate linkage to care (Tucker et al., 2015).

Our study also showed that implementation of HIVST will raise concerns related to increased risky sexual behaviours, some of them reported an increased number of multiple and concurrent partners in obtaining non-reactive results. AYP explained that unknown HIV status has been hindering them from engaging in sexual behaviours and now that they could carry out HIVST this will be an opportunity to have multiple sexual partners. Similar results were reported by Tonen-Wolyec et al. (2019) were individuals who had one HIV test had fewer sexual partners than individuals who had tested five or more times. It is important to have a non-reactive screening test before engaging in sexual activities, however, AYP must be encouraged to engage themselves in other HIV testing services to reduce their risk of acquiring HIV. The results also indicated that on testing negative, AYP are more likely not to use HIV prevention methods like condoms with their sexual partners an aspect they highlighted to increase not only the number of unplanned pregnancies but also sexually transmitted infections. Another study conducted among female sex workers also revealed similar findings where some of the sex workers stopped using condoms with their boyfriends when they obtained non-reactive self-test results (Boisvert Moreau et al., 2022).

In addition, Participants in our study believed that the introduction of self-testing services would lead to a deliberate spread of HIV. The view is that self-testing would create room for malicious AYP to spread HIV to their sexual partners after testing positive with the kit. The increased risks of HIV and STI were linked to the absence of HIV counselling services in the model. Similar results were reported in an exploratory study in Tanzania. (Nnko et al., 2020) Therefore, when rolling out HIVST among AYP, implementers must integrate it with other HIV services like HIV counselling, STI screening and treatment to help adolescents live safer and healthier lives.

While participants in this study expressed concerns about difficulties in interpreting or potentially misinterpreting the test results—especially among AYP with lower literacy levels or limited knowledge of the kit recent evidence from a systematic review on HIVST suggests that laypersons can accurately perform the test with little or no supervision from healthcare providers (Lippman et al., 2018). However, the authors of the review cautioned that accuracy in reading the test results must still be closely monitored to prevent errors, particularly among those with limited knowledge or literacy (Lippman et al., 2018). In addition, the study revealed that HIVST AYP is more likely to deny positive results on screening with the kit, this was linked to the notion that the kit may be spoilt or be reactive to any virus but not HIV.

The concern that HIVST kits were perceived as discriminatory underscores the need for more inclusive and accessible testing options that accommodate diverse populations, especially for those with visual impairments. This highlights a significant gap in the current design and distribution of the kits, suggesting that they may not fully meet the needs of all users, particularly those with disabilities or other specific conditions (Chipanta et al., 2023). Addressing this issue could inform future policy changes and influence the development of more universally accessible HIVST kits. Notably, no other studies in the existing literature have reported this concern, making this an important contribution to the discourse on HIVST. Additionally, the study revealed that many AYP expressed doubts about the accuracy of oral HIVST kits, as they believed that HIV could only be detected through blood samples. This misconception points to a need for improved education and clear communication about how different HIV testing methods work (Kurth et al., 2016; Rivera et al., 2021). These findings have significant implications for the design of HIVST interventions, as addressing both the perceived inclusivity and accuracy of the kits could enhance their acceptability and uptake among young people. However, our results agreed with other studies from Uganda that revealed concern about the accuracy of the results (Hamilton et al., 2021; Matovu et al., 2018).

The strength of the study was the views of young people were captured using a wide range of qualitative data methods. This allowed us to triangulate our results and increased the rigour of our findings. The selective sample from ongoing randomised control trials may not reflect the representativeness of a wider study population in both settings which was a limitation of this study.

## Conclusion

Despite the perceived concerns surrounding HIVST, its potential benefits and utility among AYP, who are at heightened risk of HIV but may be less likely to seek traditional healthcare services, cannot be overlooked. HIVST offers a promising avenue for reaching underserved populations, especially in settings where access to care is limited. However, to maximise the impact of HIVST, it is essential to address key concerns such as social harms, potential increases in risky behaviour, and the challenge of ensuring accurate test interpretation. Additionally, improving the utility of HIVST will require robust strategies to support linkage to essential services, including counselling, confirmatory testing, and timely access to prevention, treatment, and care services.

A successful rollout of HIVST should be accompanied by mechanisms to monitor and mitigate these potential risks, ensuring that concerns raised by policymakers and other stakeholders are effectively addressed. This includes developing clear educational materials to improve understanding of HIVST, addressing misconceptions about testing accuracy, and implementing systems for tracking adverse outcomes such as discrimination or misinterpretation of results. By doing so, HIVST can be better positioned to fulfil its promise as a key tool in HIV prevention and care. Careful evaluation of these factors will be crucial to ensuring widespread acceptance, effective utilisation, and sustained impact of HIVST in our setting. We strongly recommend that future implementation efforts prioritise the ongoing assessment of social, behavioural, and health system impacts to optimise the long-term success of this strategy.

## Data Availability

All relevant data are within the manuscript and its Supporting Information files.

## Acknowledgements

We acknowledge our study participants as well as the field mobilisers who made this study possible.

## Authors’ contributions

All authors contributed to the overall study design and specific methodologies. RM, ASS, DN and FR, were involved in data collection and analysis. MM, CB, VB, MS, JS, RN, EA, FR, JN, NJ supported the analysis, RM, ASS and DN wrote the first draft. All authors approved the final version for submission.

## References

1. Bbuye, M., Muttamba, W., Nassaka, L., Nakyomu, D., Taasi, G., Kiguli, S., . . . Mukose, A. D. (2022). Factors Associated with Linkage to HIV Care Among Oral Self-Tested HIV Positive Adults in Uganda. HIV AIDS (Auckl), 14, 61–72. doi:10.2147/hiv.S346951

2. Boisvert Moreau, M., Kintin, F. D., Atchekpe, S., Batona, G., Béhanzin, L., Guédou, F. A., . . . Alary, M. (2022). HIV self-testing implementation, distribution and use among female sex workers in Cotonou, Benin: a qualitative evaluation of acceptability and feasibility. BMC public health, 22(1), 1–13.

3. Bronfenbrenner, U. (1994). Ecological models of human development. International encyclopedia of education/Pergamon Press/Elsevier Science.

4. Brown, A. N., Djimeu, E. W., & Cameron, D. B. (2014). A review of the evidence of harm from self-tests. AIDS Behav, 18 Suppl 4(Suppl 4), S445–449. doi:10.1007/s10461-014-0831-y

5. Bryman, A. (2016). Social research methods: Oxford university press.

6. Bulterys, M. A., Naughton, B., Mujugira, A., Mugisha, J., Nakyanzi, A., Naddunga, F., . . . Sharma, M. (2023). Pregnant women and male partner perspectives of secondary distribution of HIV self-testing kits in Uganda: A qualitative study. PLoS One, 18(2), e0279781. doi:10.1371/journal.pone.0279781

7. Burke, V. M., Nakyanjo, N., Ddaaki, W., Payne, C., Hutchinson, N., Wawer, M. J., . . . Kennedy, C. E. (2017). HIV self-testing values and preferences among sex workers, fishermen, and mainland community members in Rakai, Uganda: A qualitative study. PLoS One, 12(8), e0183280. doi:10.1371/journal.pone.0183280

8. Bustamante, M. J., Konda, K. A., Joseph Davey, D., León, S. R., Calvo, G. M., Salvatierra, J., . . . Klausner, J. D. (2017). HIV self-testing in Peru: questionable availability, high acceptability but potential low linkage to care among men who have sex with men and transgender women. Int J STD AIDS, 28(2), 133–137. doi:10.1177/0956462416630674

9. Bwalya, C., Simwinga, M., Hensen, B., Gwanu, L., Hang’andu, A., Mulubwa, C., . . . Bond, V. (2020). Social response to the delivery of HIV self-testing in households: experiences from four Zambian HPTN 071 (PopART) urban communities. AIDS Res Ther, 17(1), 32. doi:10.1186/s12981-020-00287-y

10. Chanda, M. M., Ortblad, K. F., Mwale, M., Chongo, S., Kanchele, C., Kamungoma, N., . . . Oldenburg, C. E. (2017). HIV self-testing among female sex workers in Zambia: A cluster randomized controlled trial. PLoS Med, 14(11), e1002442. doi:10.1371/journal.pmed.1002442

11. Chipanta, D., Mitra, S., Amo-Agyei, S., Velarde, M. R., Amekudzi, K., Osborne, C., . . . Keiser, O. (2023). Differences between persons with and without disability in HIV prevalence, testing, treatment, and care cascade in Tanzania: a cross-sectional study using population-based data. BMC Public Health, 23(1), 2096. doi:10.1186/s12889-023-17013-8

12. Hamilton, A., Thompson, N., Choko, A. T., Hlongwa, M., Jolly, P., Korte, J. E., & Conserve, D. F. (2021). HIV Self-Testing Uptake and Intervention Strategies Among Men in Sub-Saharan Africa: A Systematic Review. Front Public Health, 9, 594298. doi:10.3389/fpubh.2021.594298

13. Hatzold, K., Gudukeya, S., Mutseta, M. N., Chilongosi, R., Nalubamba, M., Nkhoma, C., . . . Corbett, E. L. (2019). HIV self-testing: breaking the barriers to uptake of testing among men and adolescents in sub-Saharan Africa, experiences from STAR demonstration projects in Malawi, Zambia and Zimbabwe. J Int AIDS Soc, 22 Suppl 1(Suppl Suppl 1), e25244. doi:10.1002/jia2.25244

14. Hector, J., Davies, M. A., Dekker-Boersema, J., Aly, M. M., Abdalad, C. C. A., Langa, E. B. R., . . . Jefferys, L. F. (2018). Acceptability and performance of a directly assisted oral HIV self-testing intervention in adolescents in rural Mozambique. PLoS One, 13(4), e0195391. doi:10.1371/journal.pone.0195391

15. Hlongwa, M., Mashamba-Thompson, T., Makhunga, S., Muraraneza, C., & Hlongwana, K. (2020). Men’s perspectives on HIV self-testing in sub-Saharan Africa: a systematic review and meta-synthesis. BMC Public Health, 20(1), 66. doi:10.1186/s12889-020-8184-0

16. Indravudh, P. P., Sibanda, E. L., d’Elbée, M., Kumwenda, M. K., Ringwald, B., Maringwa, G., . . . Hatzold, K. (2017). ‘I will choose when to test, where I want to test’: investigating young people’s preferences for HIV self-testing in Malawi and Zimbabwe. Aids, 31, S203–S212.

17. Jamil, M. S., Eshun-Wilson, I., Witzel, T. C., Siegfried, N., Figueroa, C., Chitembo, L., . . . Johnson, C. (2021). Examining the effects of HIV self-testing compared to standard HIV testing services in the general population: A systematic review and meta-analysis. eClinicalMedicine, 38. doi:10.1016/j.eclinm.2021.100991

18. Johnson, C. C., Kennedy, C., Fonner, V., Siegfried, N., Figueroa, C., Dalal, S., . . . Baggaley, R. (2017). Examining the effects of HIV self-testing compared to standard HIV testing services: a systematic review and meta-analysis. J Int AIDS Soc, 20(1), 21594. doi:10.7448/ias.20.1.21594

19. Kalibbala, D., Mpungu, S. K., Ssuna, B., Muzeyi, W., Mberesero, H., Semitala, F. C., . . . Musiime, V. (2022). Determinants of testing for HIV among young people in Uganda. A nested, explanatory-sequential study. PLOS Glob Public Health, 2(12), e0000870. doi:10.1371/journal.pgph.0000870

20. Kaufman, M. R., Cornish, F., Zimmerman, R. S., & Johnson, B. T. (2014). Health behavior change models for HIV prevention and AIDS care: practical recommendations for a multi-level approach. Journal of acquired immune deficiency syndromes (1999), 66(Suppl 3), S250.

21. Kilanowski, J. F. P. R. A. C. F. (2017). Breadth of the Socio-Ecological Model. J Agromedicine, 22(4), 295–297. doi:10.1080/1059924x.2017.1358971

22. Kumwenda, M. K., Johnson, C. C., Choko, A. T., Lora, W., Sibande, W., Sakala, D., . . . Corbett, E. L. (2019). Exploring social harms during distribution of HIV self-testing kits using mixed-methods approaches in Malawi. J Int AIDS Soc, 22 Suppl 1(Suppl Suppl 1), e25251. doi:10.1002/jia2.25251

23. Kurth, A. E., Cleland, C. M., Chhun, N., Sidle, J. E., Were, E., Naanyu, V., . . . Siika, A. M. (2016). Accuracy and Acceptability of Oral Fluid HIV Self-Testing in a General Adult Population in Kenya. AIDS Behav, 20(4), 870–879. doi:10.1007/s10461-015-1213-9

24. Lippman, S. A., Gilmore, H. J., Lane, T., Radebe, O., Chen, Y. H., Mlotshwa, N., . . . McIntyre, J. (2018). Ability to use oral fluid and fingerstick HIV self-testing (HIVST) among South African MSM. PLoS One, 13(11), e0206849. doi:10.1371/journal.pone.0206849

25. Masters, S. H., Agot, K., Obonyo, B., Napierala Mavedzenge, S., Maman, S., & Thirumurthy, H. (2016). Promoting Partner Testing and Couples Testing through Secondary Distribution of HIV Self-Tests: A Randomized Clinical Trial. PLoS Med, 13(11), e1002166. doi:10.1371/journal.pmed.1002166

26. Matovu, J. K. B., Bogart, L. M., Nakabugo, J., Kagaayi, J., Serwadda, D., Wanyenze, R. K., . . . Kurth, A. E. (2020). Feasibility and acceptability of a pilot, peer-led HIV self-testing intervention in a hyperendemic fishing community in rural Uganda. PLoS One, 15(8), e0236141. doi:10.1371/journal.pone.0236141

27. Matovu, J. K. B., Kisa, R., Buregyeya, E., Chemusto, H., Mugerwa, S., Musoke, W., . . . Wanyenze, R. K. (2018). ’If I had not taken it [HIVST kit] home, my husband would not have come to the facility to test for HIV’: HIV self-testing perceptions, delivery strategies, and post-test experiences among pregnant women and their male partners in Central Uganda. Glob Health Action, 11(1), 1503784. doi:10.1080/16549716.2018.1503784

28. Mulubwa, C., Hensen, B., Phiri, M. M., Shanaube, K., Schaap, A. J., Floyd, S., Ayles, H. (2019). Community based distribution of oral HIV self-testing kits in Zambia: a cluster-randomised trial nested in four HPTN 071 (PopART) intervention communities. Lancet HIV, 6(2), e81–e92. doi:10.1016/s2352-3018(18)30258-3

29. Murewanhema, G., Musuka, G., Moyo, P., Moyo, E., & Dzinamarira, T. (2022). HIV and adolescent girls and young women in sub-Saharan Africa: A call for expedited action to reduce new infections. IJID Reg, 5, 30–32. doi:10.1016/j.ijregi.2022.08.009

30. Napierala Mavedzenge, S., Baggaley, R., & Corbett, E. L. (2013). A review of self-testing for HIV: research and policy priorities in a new era of HIV prevention. Clin Infect Dis, 57(1), 126–138. doi:10.1093/cid/cit156

31. Nasuuna, E., Namimbi, F., Muwanguzi, P. A., Kabatesi, D., Apolot, M., Muganzi, A., & Kigozi, J. (2022). Early observations from the HIV self-testing program among key populations and sexual partners of pregnant mothers in Kampala, Uganda: A cross sectional study. PLOS Global Public Health, 2(1), e0000120.

32. Naughton, B., Bulterys, M. A., Mugisha, J., Mujugira, A., Boyer, J., Celum, C., . . . Sharma, M. (2023). ’If there is joy… I think it can work well’: a qualitative study investigating relationship factors impacting HIV self-testing acceptability among pregnant women and male partners in Uganda. BMJ Open, 13(2), e067172. doi:10.1136/bmjopen-2022-067172

33. Njau, B., Lisasi, E., Damian, D. J., Mushi, D. L., Boulle, A., & Mathews, C. (2020). Feasibility of an HIV self-testing intervention: a formative qualitative study among individuals, community leaders, and HIV testing experts in northern Tanzania. BMC Public Health, 20(1), 490. doi:10.1186/s12889-020-08651-3

34. Nnko, S., Nyato, D., Kuringe, E., Casalini, C., Shao, A., Komba, A., . . . Wambura, M. (2020). Female sex workers perspectives and concerns regarding HIV self-testing: an exploratory study in Tanzania. BMC Public Health, 20(1), 1–9.

35. Nsereko, G. M., Kobusingye, L. K., Musanje, K., Nangendo, J., Nantamu, S., & Baluku, M. M. (2024). Self-testing knowledge and beliefs on HIV self-testing use in central Uganda. PLOS Glob Public Health, 4(6), e0002869. doi:10.1371/journal.pgph.0002869

36. Obiezu-Umeh, C., Gbajabiamila, T., Ezechi, O., Nwaozuru, U., Ong, J. J., Idigbe, I., . . . Iwelunmor, J. (2021). Young people’s preferences for HIV self-testing services in Nigeria: a qualitative analysis. BMC Public Health, 21(1), 67. doi:10.1186/s12889-020-10072-1

37. Ortblad, K., Kibuuka Musoke, D., Ngabirano, T., Nakitende, A., Harling, G., Haberer, J. E., . . . Bärnighausen, T. (2019). The Effect of HIV Self-Testing Delivery Models on Female Sex Workers’ Sexual Behaviors: A Randomized Controlled Trial in Urban Uganda. AIDS Behav, 23(5), 1225–1239. doi:10.1007/s10461-019-02393-z

38. Ortblad, K., Kibuuka Musoke, D., Ngabirano, T., Nakitende, A., Magoola, J., Kayiira, P., . . . Bärnighausen, T. (2017). Direct provision versus facility collection of HIV self-tests among female sex workers in Uganda: A cluster-randomized controlled health systems trial. PLoS Med, 14(11), e1002458. doi:10.1371/journal.pmed.1002458

39. Phongphiew, P., Songtaweesin, W. N., Paiboon, N., Phiphatkhunarnon, P., Srimuan, P., Sowaprux, T., . . . Puthanakit, T. (2021). Acceptability of blood-based HIV self-testing among adolescents aged 15-19 years at risk of HIV acquisition in Bangkok. Int J STD AIDS, 32(10), 927–932. doi:10.1177/09564624211003742

40. Rivera, A. S., Hernandez, R., Mag-Usara, R., Sy, K. N., Ulitin, A. R., O’Dwyer, L. C., . . . Hirschhorn, L. R. (2021). Implementation outcomes of HIV self-testing in low- and middle- income countries: A scoping review. PLoS One, 16(5), e0250434. doi:10.1371/journal.pone.0250434

41. Schaffer, E. M., Agot, K., & Thirumurthy, H. (2017). The Association Between Intimate Partner Violence and Women’s Distribution and Use of HIV Self-Tests With Male Partners: Evidence From a Cohort Study in Kenya. J Acquir Immune Defic Syndr, 76(3), e85–e87. doi:10.1097/qai.0000000000001502

42. Sharma, M., Ying, R., Tarr, G., & Barnabas, R. (2015). Systematic review and meta-analysis of community and facility-based HIV testing to address linkage to care gaps in sub-Saharan Africa. Nature, 528(7580), S77–85. doi:10.1038/nature16044

43. Sileo, K. M., Kintu, M., Chanes-Mora, P., & Kiene, S. M. (2016). “Such Behaviors Are Not in My Home Village, I Got Them Here”: A Qualitative Study of the Influence of Contextual Factors on Alcohol and HIV Risk Behaviors in a Fishing Community on Lake Victoria, Uganda. AIDS Behav, 20(3), 537–547. doi:10.1007/s10461-015-1077-z

44. Spielberg, F., Levine, R. O., & Weaver, M. (2004). Self-testing for HIV: a new option for HIV prevention? Lancet Infect Dis, 4(10), 640–646. doi:10.1016/s1473-3099(04)01150-8

45. Tonen-Wolyec, S., Batina-Agasa, S., Muwonga, J., Mboumba Bouassa, R. S., Kayembe Tshilumba, C., & Bélec, L. (2019). Acceptability, feasibility, and individual preferences of blood-based HIV self-testing in a population-based sample of adolescents in Kisangani, Democratic Republic of the Congo. PLoS One, 14(7), e0218795. doi:10.1371/journal.pone.0218795

46. Tucker, J. D., Wei, C., Pendse, R., & Lo, Y. R. (2015). HIV self-testing among key populations: an implementation science approach to evaluating self-testing. J Virus Erad, 1(1), 38–42.

47. UNAIDS. (2021). Update. Confronting Inequalities-Lessons for pandemic responses from 40 years of AIDS. Retrieved from UNAIDS. (2025). UNAIDS 2025 epidemiological estimates. Retrieved from https://www.unaids.org/sites/default/files/media_asset/UNAIDS_FactSheet_en.pdf

48. UNAIDS, A. (2014). A short technical update on self-testing for HIV. Geneva: UNAIDS, WHO.

49. van Sighem, A., & van der Valk, M. (2022). Moving towards zero new HIV infections: The importance of combination prevention. Lancet Reg Health West Pac, 25, 100558. doi:10.1016/j.lanwpc.2022.100558

50. Wilson, K. S., Mugo, C., Katz, D. A., Manyeki, V., Mungwala, C., Otiso, L., . . . Kohler, P. K. (2022). High Acceptance and Completion of HIV Self-testing Among Diverse Populations of Young People in Kenya Using a Community-Based Distribution Strategy. AIDS Behav, 26(3), 964–974. doi:10.1007/s10461-021-03451-1

51. World Health Organisation. (2019). WHO HIV Policy and Implementation status in Countries. Retrieved from Geneva: https://iris.who.int/bitstream/handle/10665/326035/WHO-CDS-HIV-19.20-eng.pdf

52. Zanolini, A., Chipungu, J., Vinikoor, M. J., Bosomprah, S., Mafwenko, M., Holmes, C. B., & Thirumurthy, H. (2018). HIV Self-Testing in Lusaka Province, Zambia: Acceptability, Comprehension of Testing Instructions, and Individual Preferences for Self-Test Kit Distribution in a Population-Based Sample of Adolescents and Adults. AIDS Res Hum Retroviruses, 34(3), 254–260. doi:10.1089/aid.2017.0156

53. Zeleke, E. A., Stephens, J. H., Gesesew, H. A., Gello, B. M., & Ziersch, A. (2024). Acceptability and use of HIV self-testing among young people in sub-Saharan Africa: a mixed methods systematic review. BMC Prim Care, 25(1), 369. doi:10.1186/s12875-024-02612-0

